# Mild cognitive impairment cases affect the predictive power of Alzheimer’s disease diagnostic models using routine clinical variables

**DOI:** 10.1101/2025.02.04.25321694

**Authors:** Caitlin A. Finney, Alzheimer’s Disease Neuroimaging Initiative, Artur Shvetcov

## Abstract

Diagnostic models using primary care routine clinical variables have been limited in their ability to identify Alzheimer’s disease (AD) patients. In this study we sought to better understand the effect of mild cognitive impairment (MCI) on the predictive performance of AD diagnostic models. We sourced data from the Alzheimer’s Disease Neuroimaging Initiative (ADNI) cohort. CatBoost was used to assess the utility of routine clinical variables that are accessible to primary care physicians, such as hematological and blood tests and medical history, in multiclass classification between healthy controls, MCI, and AD. Our results indicated that MCI indeed affected the predictive performance of AD diagnostic models. Of three subgroups of MCI that we found, this finding was driven by a subgroup of MCI patients that likely have prodromal AD. Future research should focus on distinguishing MCI from prodromal AD as the utmost priority for improving translational AD diagnostic models for primary care physicians.

## Introduction

Alzheimer’s disease (AD), the most prevalent dementia accounting for 50-70% of cases, is the leading cause of disability among adults over age 65 ^1^. With a rapidly increasing prevalence, AD is expected to cost the world economy more than $14.5 trillion international dollars over the next 30 years to 2050 ^2^. Improving the ability to accurately diagnose AD is of the utmost importance and ensures that patients receive appropriate support, interventions, and have the time to employ lifestyle adjustments that prolong independence and quality of life ^3^.

Despite this, the timely and accurate diagnosis of AD remains challenging. To date, a significant number of diagnostic tools and models of AD have focused on the use of magnetic resonance imaging (MRI) and positron emission tomography (PET) scans to detect early indicators of AD pathology including Aβ plaques ^4–12^. Others have relied on cerebrospinal fluid (CSF) and plasma biomarkers including Aβ_42_/Aβ_40_ ratio, total tau protein, phosphorylated tau 181 (p-tau_181_), p-tau_231_, p-tau_217_, neurofilament light (NfL) and glial fibrillary acidic protein (GFAP) ^13,14^. Although these tests show promise for AD diagnostics, there are many practical limitations that prevent widespread clinical implementation. Both neuroimaging scans and biomarker assays are associated with a high cost both to the healthcare system and patient, low availability, and high wait times, especially for those patients in rural areas ^14–17^. Further, collecting CSF is an invasive procedure that requires specific technical medical expertise ^14^. In line with these challenges, a recent Alzheimer’s Association Primary Care Physician Dementia Care Training Survey found that half of primary care physicians do not feel that they have the local specialist resources to meet patient demand ^18^. In fact, primary care physicians remain better able to identify those without AD than those with it ^19,20^. For many patients with AD, primary care physicians are the first point of contact with the healthcare system making them essential for patient triage, diagnosis, and management ^15^. Therefore, ensuring that primary care physicians have the skills and tools required for AD diagnostics is critical.

To improve diagnostic capabilities among primary care physicians, previous studies have used machine learning to examine the potential of routine, easy-to-obtain clinical measures. For example, these models include the Cardiovascular Risk Factors, Aging, and Dementia (CAIDE) ^21^, Study on Aging, Cognition and Dementia (AgeCoDe) ^22^, Australian National University Alzheimer’s Disease Risk Index (ANU-ADRI) ^23^, Rapid Assessment of Dementia Risk (RADaR) for older adults ^24^, and Brief Dementia Screening Indicator (BDSI)^25^. There are limitations to these previous diagnostic models, however, as they report low sensitivities and positive predictive values (PPV), indicating that they are unable to reliably identify someone with AD. This was confirmed by a recent study showing existing machine learning-based diagnostic models of AD miss 84-91% of incident AD cases and therefore have limited clinical utility ^26^.

The factors underlying the inability of these models to reliably diagnose AD remains unclear. One possibility is the presence of patients with mild cognitive impairment (MCI), which affects 10-15% of the population over age 65 ^27^. Although AD first manifests clinically as MCI, not all patients with MCI will go on to develop AD ^27–29^. Further, MCI is known to be characterized by heterogeneous patients with multiple etiologies associated with differing clinical presentations ^28^. In line with this, there is evidence that MCI cases themselves are difficult to predict. Previous studies have reported precision, recall, and sensitivity metrics at or near chance levels for MCI classification ^30,31^, distinguishing MCI from AD ^32^, and predicting conversion from MCI to AD ^33–36^. This suggests that these models would have low practical utility in the clinic and are likely to mislabel most MCI cases. Combined, this highlights that MCI cases may affect the predictive power of translational AD diagnostic models.

Leveraging data from the Alzheimer’s Disease Neuroimaging Initiative (ADNI) cohort, we sought to better understand the effects of MCI cases on the predictive performance of AD diagnostic models using easy-to-obtain clinical variables.

## Results

### CatBoost can identify healthy controls and MCI but not AD cases

Using baseline diagnosis, we identified patients in the ADNI cohort with AD (N = 181), MCI (N = 473), and healthy controls (N = 220). There were no differences across the groups with respect to age or sex distributions, with all groups having more males than females (Supplementary Table 1). The dataset was then randomly split into 80% training and validation and 20% withheld testing datasets. We included 120 features of routine, easy-to-obtain clinical variables (Supplementary Table 1). Using CatBoost, we first sought to determine whether any of these features were able to differentiate between healthy control, MCI, and AD patients. Our multiclass classification model was able to successfully identify healthy controls, showing high performance metrics ≥ 0.82 (Table 1). Performance metrics for MCI identification were lower, however, at ≥ 0.75 (Table 1). In the case of healthy controls, performance metrics suggested that the models were largely unable to successfully differentiate AD cases, with a very low sensitivity of 0.63 and PPV of 0.65 (Table 1).

**Table 1.** Performance metrics of a CatBoost model for diagnosing healthy controls, MCI, and AD using 120 clinical variables as features.

|  | **Sensitivity** | **Specificity** | **PPV** | **NPV** | **AUC** |
| --- | --- | --- | --- | --- | --- |
| Healthy control | 0.93 | 0.93 | 0.82 | 0.98 | 0.98 |
| MCI patients | 0.78 | 0.79 | 0.83 | 0.75 | 0.86 |
| AD patients | 0.63 | 0.91 | 0.65 | 0.91 | 0.90 |

Given that our model was able to successfully identify healthy control cases, we hypothesized that it was having difficulty differentiating between MCI and AD cases, specifically. A confusion matrix showed that this was indeed the case (Figure 1A). The model only misclassified 9 healthy controls as MCI patients. This increased, however, to a misclassification of 13 MCI cases as AD and 12 AD cases as MCI (Figure 1A).

**Figure 1.**
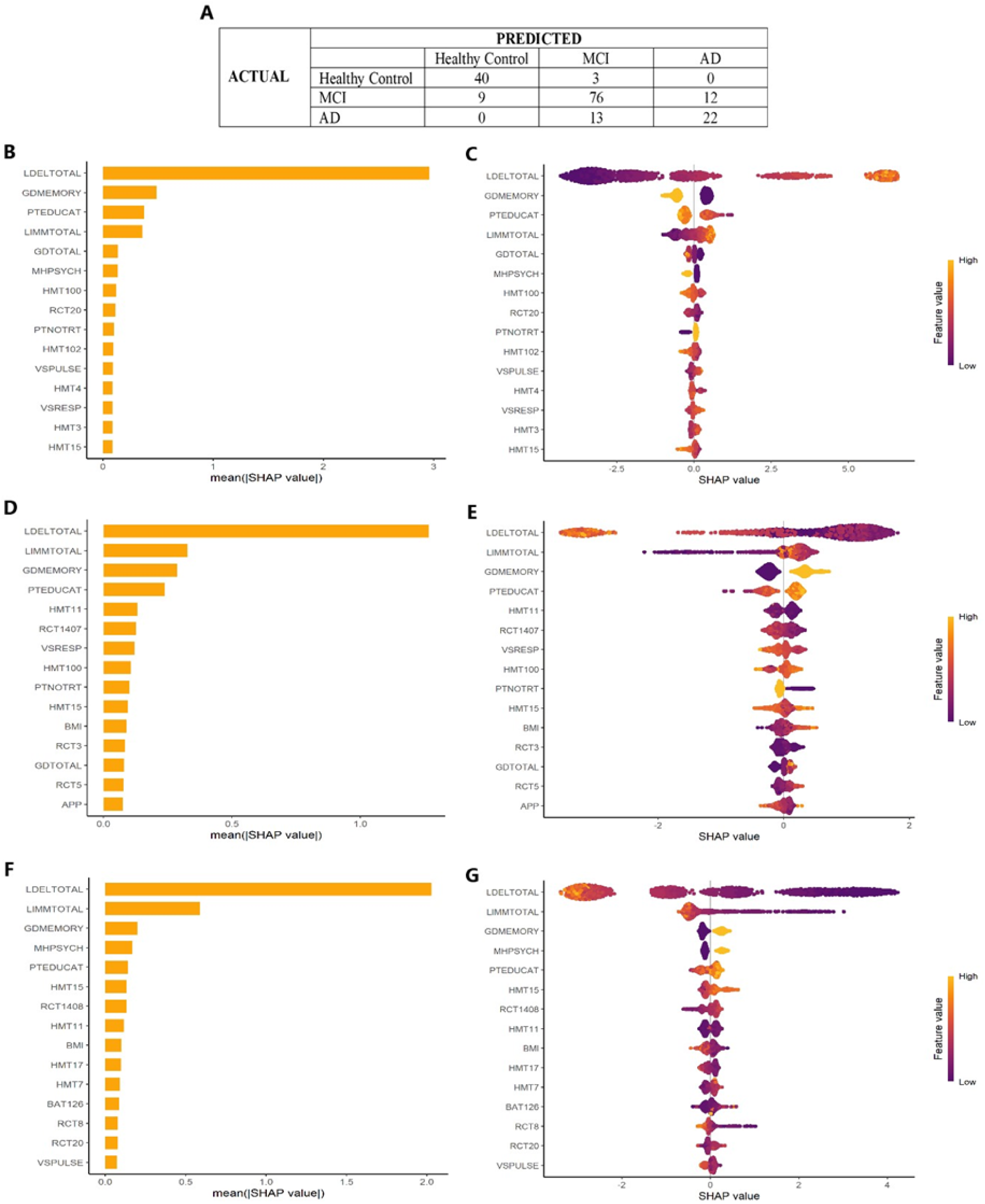
Performance of a CatBoost model and feature importance for diagnosis of healthy control, MCI, and AD. (**A**) Confusion matrix showing the true (correct) and false (incorrect) prediction of healthy control, MCI, and AD. (**B**) Absolute SHAP values and (**C**) heat map of the contribution of the main features for predicting healthy controls. (**D**) Absolute SHAP values and (**E**) heat map of the contribution of the main features for predicting MCI. (**F**) Absolute SHAP values and (**G**) heat map of the contribution of the main features for predicting AD. Abbreviations: APP: augmented pulse pressure; BAT126: vitamin B12; BMI: body mass index; GDMEMORY: item 10 of Geriatric Depression Scale; GDTOTAL: total score Geriatric Depression Scale; HMT3: red blood cell count; HMT4: mean corpuscular volume; HMT7: white blood cell count; HMT11: eosinophils; HMT15: neutrophils; HMT17: white blood cell count; HMT100: mean corpuscular hemoglobin; HMT102: mean corpuscular hemoglobin concentration; LDELTOTAL: total number of story units recalled on Logical Memory-Delayed Recall; LIMMTOTAL: total number of story units recalled on Logical Memory-Immediate Recall; MHPSYCH: psychiatric medical history; PTEDUCAT: education; PTNOTRT: retirement; RCT3: gamma-glutamyl transferase; RCT: aspartate aminotransferase (serum glutamic-oxaloacetic transaminase); RCT8: serum uric acid; RCT20: cholesterol; RCT1407: alkaline phosphatase; RCT1408: lactate dehydrogenase; VSPULSE: seated pulse rate (per minute); VSRESP: respirations (per minute).

To better understand which features were being used by our CatBoost model to predict between the groups, we performed a SHAP analysis. The five most important features for identifying healthy controls included the Logical Memory-Delayed Recall, item 10 of the Geriatric Depression Scale that asks about memory problems, years of education, Logical Memory-Immediate Recall, and total score on the Geriatric Depression Scale (Figure 1B,C). For MCI cases, the total score on the Geriatric Depression Scale was replaced by eosinophils (Figure 1D,E) and by psychiatric medical history for AD (Figure 1F,G).

### Feature selection slightly improves the predictive performance of CatBoost for identifying MCI and AD patients

To identify if we could improve the performance of CatBoost model, we performed filter-based feature selection to include only those variables significantly associated with the outcome (diagnosis). This resulted in 19 features being included (Supplementary Table 2).

As before, our model was successfully able to identify healthy controls, with only a small reduction in PPV (Table 2). There was also a negligible reduction, relative to the model with 120 features, in the predictive performance for identifying MCI cases (Table 2). For AD cases, however, filter-based feature selection improved the predictive performance, with sensitivity increasing from 0.63 to 0.74 (Table 2).

**Table 2.** Performance metrics of a CatBoost model for diagnosing healthy controls, MCI, and AD following feature selection.

|  | Sensitivity | Specificity | PPV | NPV | AUC |
| --- | --- | --- | --- | --- | --- |
| Healthy control | 0.93 | 0.92 | 0.78 | 0.98 | 0.98 |
| MCI patients | 0.71 | 0.85 | 0.85 | 0.70 | 0.90 |
| AD patients | 0.74 | 0.88 | 0.60 | 0.93 | 0.93 |

These changes in predictive performance were also reflected in the confusion matrix. Here, 11 healthy controls were misclassified as MCI whereas only 9 cases of MCI were misclassified as AD and 17 AD cases misclassified as MCI (Figure 2A). This resulted in a significant drop in PPV of AD cases from 0.65 down to 0.60 here.

**Figure 2.**
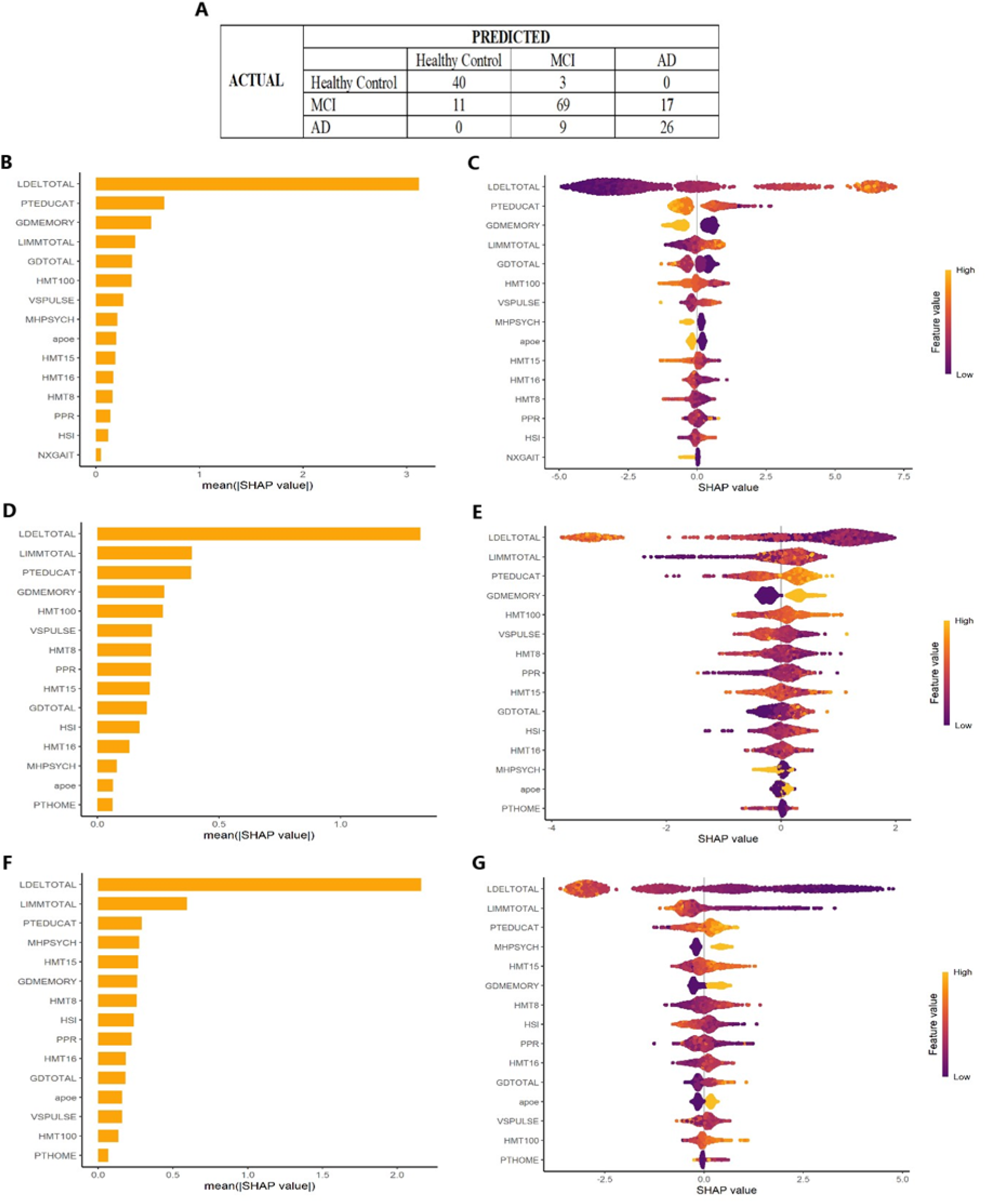
Performance of the CatBoost model and feature importance for diagnosis of healthy control, MCI, and AD after filter-based feature selection. (**A**) Confusion matrix showing the true (correct) and false (incorrect) prediction of healthy control, MCI, and AD. (**B**) Absolute SHAP values and (**C**) heat map of the contribution of the main features for predicting healthy controls. (**D**) Absolute SHAP values and (**E**) heat map of the contribution of the main features for predicting MCI. (**F**) Absolute SHAP values and (**G**) heat map of the contribution of the main features for predicting AD. Abbreviations: apoe: apolipoprotein E genotype; GDMEMORY: item 10 of Geriatric Depression Scale; GDTOTAL: total score Geriatric Depression Scale; HMT8: neutrophils; HMT15: percent neutrophils; HMT16: lymphocytes; HMT100: mean corpuscular hemoglobin; HSI: heart stress index; LDELTOTAL: total number of story units recalled on Logical Memory-Delayed Recall; LIMMTOTAL: total number of story units recalled on Logical Memory-Immediate Recall; MHPSYCH: psychiatric medical history; NXGAIT: gait on neurological exam; PPR: pulse to pressure ratio; PTEDUCAT: education; VSPULSE: seated pulse rate (per minute).

Across healthy controls, MCI, and AD, a SHAP analysis indicated that the most important features for predicting all three groups were Logical Memory-Delayed Recall, Logical Memory-Immediate Recall, and years of education, (Figure 2B-G). For healthy controls, additional features included memory-related item 10 and total score of the Geriatric Depression Scale (Figure 2B-C). For MCI, item 10 of the Geriatric Depression Scale was also important and mean corpuscular hemoglobin (Figure 2D-E). The two additional features that were important for predicting AD patients were psychiatric medical history and percent neutrophils.

### Poor predictive performance of CatBoost models is driven by a subgroup of MCI patients that are characteristically similar to those with AD

Given that MCI is a made up of a highly heterogenous group of patients ^27–29^, we hypothesized that MCI patients may be affecting the predictive performance of AD diagnostic models. To identify if this was the case, we first tested the ability of CatBoost to distinguish between only healthy control and AD patients using 19 features previously identified using filter-based feature selection. Reducing our model to a simple binary classification resulted in the ability to readily distinguish between healthy control and AD patients as indicated by high performance metrics (>0.98; Table 3).

**Table 3.** Performance metrics of CatBoost for diagnosing healthy control or AD patients.

|  | **Sensitivity** | **Specificity** | **PPV** | **NPV** | **AUC** |
| --- | --- | --- | --- | --- | --- |
| All features | 1.00 | 1.00 | 1.00 | 1.00 | 1.00 |
| Selected features | 1.00 | 1.00 | 1.00 | 1.00 | 1.00 |

We next sought to understand why MCI patients were affecting predictive performance of our diagnostic models. Using a principal component analysis (PCA) based on our 19 identified features, we found that while there was clear group separation between healthy control and AD patients MCI patients were distributed across both clusters (Figure 3A). We identified that the MCI group could be divided into three distinct subclusters: one that overlapped healthy controls (MCI-Healthy), one that significantly overlapped with AD patients (MCI-AD), and a third that was distinct yet more closely related to AD (MCI-MCI; Figure 3B). This was further confirmed by a hierarchical cluster dendrogram (Figure 3C) and inertia plot (Figure 3D).

**Figure 3.**
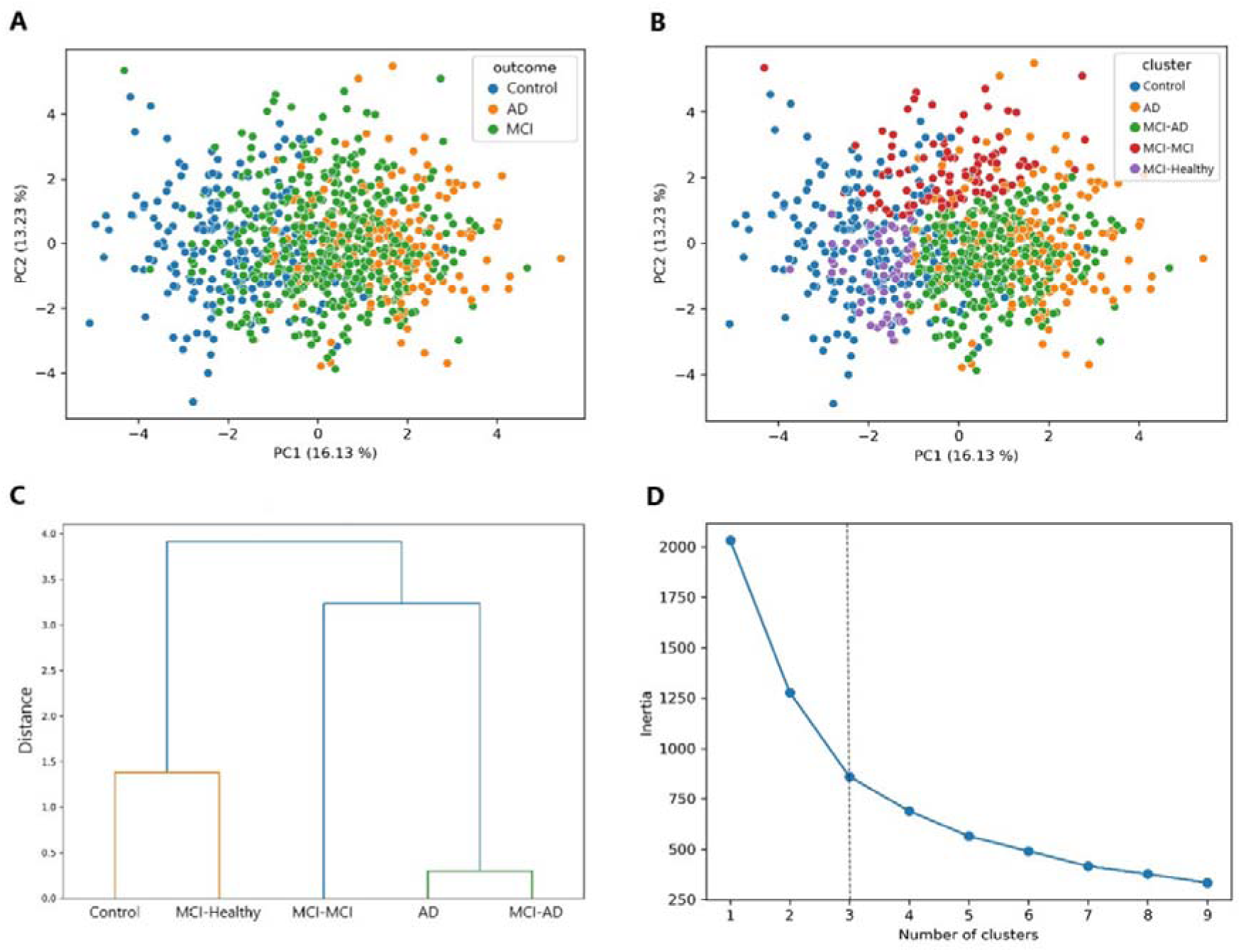
Identification of the distinct subgroups of MCI patients. (**A**) Principal component analysis showing clear separation between healthy control and AD patients but significant overlap of MCI. (**B**) Principal component analysis showing that MCI forms three distinct subclusters (MCI-Healthy, MCI-AD, and MCI-MCI) that have varying amounts of overlap with healthy control and AD. (**C**) Hierarchical cluster dendrogram showing that MCI-Healthy is closely related to healthy control, MCI-AD is closely related to AD and that MCI-MCI is distinct but more related to AD. (**D**) Elbow plot showing the relationship between the number of clusters and the within-cluster sum of squared distances (inertia). The dotted line represents the optimal number of clusters.

We then looked to characterize the three subgroups of MCI using the top significantly different features (see Supplementary Table 3 for statistical analyses). This showed that the three MCI subgroups differed on apolipoprotein E ε4 (APOE4) genotype (Figure 4A), lymphocyte count (Figure 4B), neutrophils (Figure 4C-D), seated pulse rate (Figure 4E), Geriatric Depression Scale item 10 on memory complaints (Figure 4F), and their total scores on the Logical Memory Immediate Recall (Figure 4G) and Delayed Recall (Figure 4H) tests. We found that MCI-Healthy was the most distinct group across these features relative to MCI-MCI and MCI-AD. The MCI-Healthy Group had a lower number of people with at least one APOE4 allele, neutrophils, and percent of people that replied yes to having memory complaints on the Geriatric Depression Scale. They also had higher lymphocytes and total scores on the Logical Memory tests.

**Figure 4.**
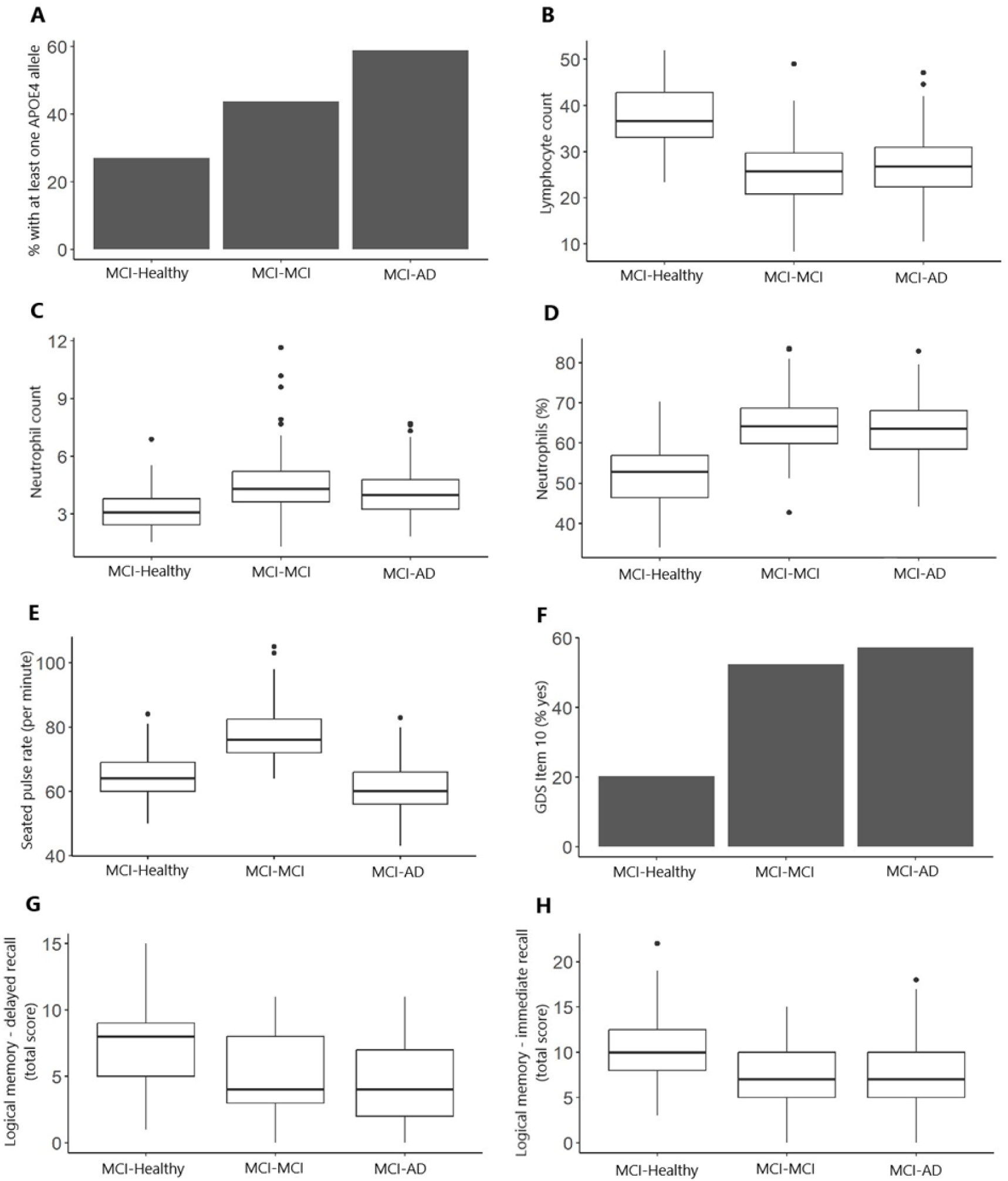
Comparison between the top significantly (*p* < 0.001) different variables between the three MCI groups (MCI-Healthy, MCI-MCI, and MCI-AD). (**A**) Percent of patients with at least one APOE4 allele. (**B**) Lymphocyte count in blood. (**C**) Neutrophil count in blood. (**D**) Percent of neutrophils in blood. (**E**) Seated pulse rate per minute. (**F**) Percent of patients that replied yes to Geriatric Depression Scale item 10 (Do you feel you have more problems with memory than most?). (**G**) Total score on the Logical Memory Delayed Recall test. (**H**) Total score on the Logical Memory Immediate Recall test.

There were also differences across the three MCI subgroups with how stable their cognitive diagnosis was or whether it changed over time. The MCI-Healthy subgroup was the most likely to maintain a stable MCI state (Figure 5A) or revert to being cognitively healthy (Figure 5B) over time. The MCI-MCI subgroup was slightly more likely than the MCI-AD subgroup to maintain MCI (Figure 5A) or revert to healthy (Figure 5B). The MCI-Healthy subgroup relative to both the MCI-MCI (X^2^ = 13.294, *p* = 0.0039) and MCI-AD groups (X^2^ = 36.483, *p* < 0.0001) was also less likely to progress to AD. There were no significant differences in the rate of progression to AD between MCI-MCI and MCI-AD subgroups (Figure 5C).

**Figure 5.**
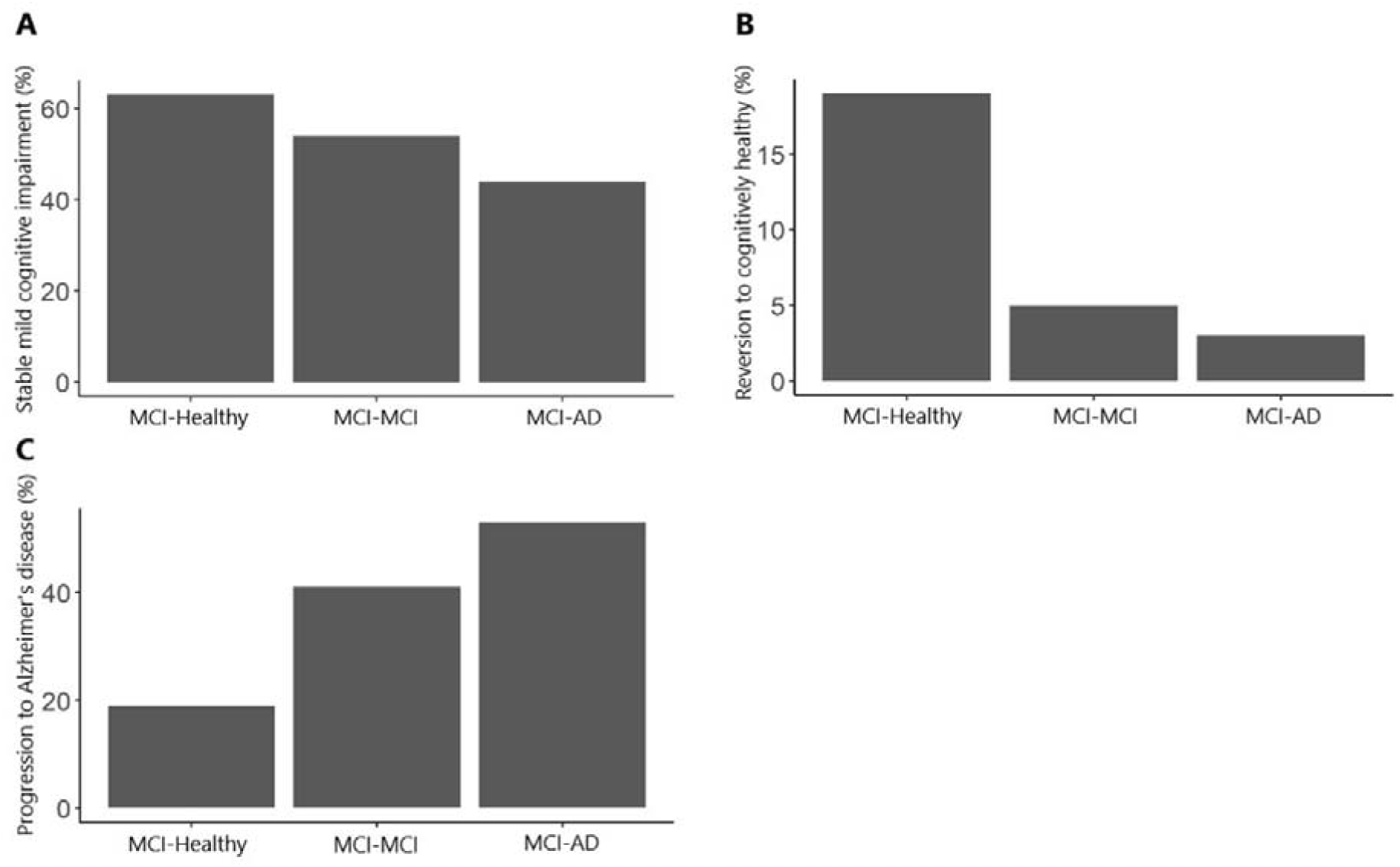
Percent of MCI patients with cognitive diagnosis stability or change over time across the three subgroups: MCI-Healthy, MCI-MCI, and MCI-AD. (**A**) Percent of MCI patients with a stable diagnosis of MCI over time. (**B**) Percent of MCI patients who revert to cognitively healthy status over time. (**C**) Percent of MCI patients who progress to AD over time.

Finally, we sought to identify which, if any, of the three MCI subgroups was driving poor predictive performance. To do this, we implemented three CatBoost models where one of the three MCI subgroups was removed and determined the models’ ability to identify healthy controls, MCI, and AD patients. When we removed either the MCI-Healthy or MCI-MCI groups, our CatBoost models were able to identify healthy controls, as before, but were still unable to identify AD patients (Table 4). When we removed the MCI-AD subgroup, however, our CatBoost model was able to identify all patient types (Table 4), suggesting that the MCI-AD subgroup, specifically, drives poor predictive performance of our diagnostic models.

**Table 4.** Performance metrics of CatBoost for diagnosing healthy control, MCI, or AD patients after removing specific MCI subgroups.

|  | **Sensitivity** | **Specificity** | **PPV** | **NPV** | **AUC** |
| --- | --- | --- | --- | --- | --- |
| **MCI-Healthy Group Removed** |  |  |  |  |  |
| Healthy control | 0.78 | 0.99 | 0.96 | 0.95 | 0.97 |
| MCI patients | 0.83 | 0.66 | 0.76 | 0.75 | 0.79 |
| AD patients | 0.56 | 0.88 | 0.59 | 0.87 | 0.84 |
| **MCI-MCI Group Removed** |  |  |  |  |  |
| Healthy control | 0.92 | 0.97 | 0.92 | 0.97 | 0.99 |
| MCI patients | 0.77 | 0.78 | 0.76 | 0.79 | 0.85 |
| AD patients | 0.64 | 0.88 | 0.66 | 0.87 | 0.86 |
| **MCI-AD Group Removed** |  |  |  |  |  |
| Healthy control | 0.93 | 0.89 | 0.86 | 0.95 | 0.96 |
| MCI patients | 0.72 | 0.92 | 0.81 | 0.87 | 0.91 |
| AD patients | 0.90 | 0.96 | 0.90 | 0.96 | 0.98 |

## Discussion

There is an urgent need to better support primary care physicians in their clinical decision making on MCI and AD. To do this, we leveraged data from the ADNI cohort and used machine learning to determine the diagnostic potential of 120 easy-to-obtain clinical measures for MCI and AD.

Using all 120 measures as features, we found that while our model could readily identify healthy control cases, it was unable to identify MCI and AD patients and had a high level of confusion between these two diagnostic categories. Using filter-based feature selection, we narrowed down the measures to 19 that were highly correlated with the outcome. Although this led to a degree of improvement in the ability to diagnose MCI and AD sensitivities were still low, ranging between 0.71 to 0.74. This suggests that our model would likely miss around 25% of MCI and AD cases. Of note, however, was that our model outperformed existing ones ^21–25,30–32^ and we therefore sought to identify the features that were most important for diagnostic prediction. Using SHAP analysis, we showed that important features included Logical Memory Delayed and Immediate Recall scores, years of education, responses to item 10 (memory) and total score on the Geriatric Depression Scale, eosinophils, and psychiatric medical history.

These findings are in line with previous research. Higher Geriatric Depression Scale scores are associated with faster cognitive decline and increased risk of MCI and AD ^37–41^. In line with this, AD and MCI patients have been shown to have increased incidence of psychiatric issues including depressive, apathy, and anxiety disorders ^42–44^. Low levels of education are also known to be associated with an increased risk of MCI and AD ^45,46^.

Peripheral neutrophil activation and lymphocytes, as well as a neutrophil-to-lymphocyte ratio, have been widely implicated in MCI and AD ^47–52^. Lower levels of hemoglobin in blood have also been linked to decreased cognitive function and AD ^52,53^. Although many studies show the importance of Logical Memory Immediate and Delayed Recall test scores for identifying healthy controls, MCI, and AD patients ^54,55^, others have indicated that they have a limited diagnostic accuracy ^56^. The reasons for these different findings are not clear and warrant further research.

An important finding of our study was that our models were confusing MCI and AD cases, specifically. To better understand why this was the case, we showed that MCI could be divided into three distinct subgroups: a subgroup that overlapped with healthy controls (MCI-Healthy), a subgroup that overlapped with AD (MCI-AD), and a subgroup that fell in between but was more closely related to AD (MCI-MCI). When we characterized these three subgroups, we found that they differed on key measures including APOE genotype, lymphocytes and neutrophils, seated pulse rate, and memory as measured by item 10 of the Geriatric Depression Scale and the Logical Memory Delayed and Immediate Recall tests. To date, there has been little consensus in the literature about how many subtypes of MCI exist^57^. In our study, we showed that in the ADNI cohort there are three distinct subgroups. Other studies, however, report between two and four to five subtypes ranging from amnesic to non-amnesic MCI ^28,58–61^. The discrepancies may lie between using *a priori* definitions of MCI, largely based on the number and type of cognitive domains that are impaired, versus our approach of data-driven *post hoc* definitions. Although a complete comparative assessment across both approaches was outside of the scope of our current study, and lack of sufficiently powered available data, future research would benefit from examining the merits and pitfalls of both.

We also found significant differences in the long-term trajectories of patients within each of the three MCI subgroups. MCI-AD patients were less likely to maintain a stable MCI diagnosis or to revert to being cognitively healthy relative to the MCI-Healthy and MCI-MCI subgroups. They were also more likely to progress to AD over time. Our MCI-AD group appears to have overlap with amnesic MCI previously reported in the literature. For example, studies show that patients with amnesic MCI are more likely to progress to AD over time ^62–64^, overlapping with our findings here for MCI-AD. Risk of amnesic MCI also increases in patients with at least one copy of the APOE ε4 genetic variant ^60^, in line with our finding that MCI-AD subgroup has the highest percentage of patients with this APOE genotype. Further, MCI-AD may be indicative of prodromal AD, as impaired delayed recall is reported to be the most early cognitive change in this group ^29^.

Interestingly, we showed that the MCI-AD group, specifically, largely drove our models’ low ability to distinguish between MCI and AD. This finding has important implications because it suggests that only one subtype of MCI leads to poor performance of diagnostic AD models. Based on the data presented here, it’s not clear how to get around this issue in practice. One solution is to further characterize these three subtypes of MCI and develop models that can distinguish between them as well as AD and healthy control cases. To do this, however, there is a need to prioritize the establishment of substantially larger, well-defined (i.e. many clinical measures obtained) cohorts of patients with MCI and to follow their trajectories over time. This is an especially important consideration in the context of supporting primary care physicians’ ability to undertake diagnostics. Primary care physicians have particular difficulty in correctly identifying MCI cases in their patients and is exacerbated by inadequate infrastructure, resources, and equipment ^15,65,66^. Current diagnostic methods for MCI largely rely on clinical judgement include subjective or objective cognitive impairment ^28,29,67^ with a preservation of basic daily functioning ^28,29^, which distinguishes it from AD. However, in practice, there appears to be limitations to clinical judgement. A recent study of Medicare data from the US representing >54,000 practices and >226,000 primary care physicians showed that only 0.1% of physicians and practices have MCI diagnosis rates within the expected range ^68^. Overall, our work highlights the importance of continuing to focus on differentiating MCI cases from prodromal AD to improve the translatability of effective diagnostic models of AD that can be used by primary care physicians in the clinic.

There are some additional considerations with respect to our findings and implementing them in practice. The first is whether primary care physicians are motivated to diagnose MCI, as some previous studies have shown that there is a low motivation due to a perceived lack of benefits for the patients ^66^. This is likely driven by a lack of effective treatments that physicians can offer their patients. It is still important, however, to acknowledge that providing an MCI, or even an MCI subtype, diagnosis, primary care physicians can provide patients and their families with assurances that what they are experiencing has a name and give them the knowledge required to plan for the future ^69^. A second consideration of our work is that primary care physicians have reported feeling that they do not have sufficient time during a brief consultation to perform broad cognitive assessments ^66^. Further some feel that they lack the neuropsychological training needed to complete this type of testing, which may limit widespread translational into the clinic ^15,67^. Our results suggest that rather than needing to learn many complex neuropsychological questionnaires, primary care physicians only need to use the Logical Memory Immediate and Delayed Recall tests and the Geriatric Depression Scale. A final limitation of our work is that we did not examine the diagnostic predictive capabilities of any AD biomarkers. For example, previous studies have linked MCI subtypes with changes in cerebrospinal fluid (CSF) total tau ^59^ and phospho-tau181 ^70^. The exclusion of this data, however, was deliberate on our part as biomarkers from plasma and CSF are still not widely used, nor are recommended for use, in clinical practice especially in the context of MCI ^15,71^. Further, primary care physicians lack the specialized equipment and expertise to routinely collect CSF samples from patients and AD biomarker panels remain expensive. Despite this, as AD biomarkers increase in popularity and become more widely available, future work would benefit from examining the diagnostic potential of these in the context of differentiating between subtypes of MCI, AD, and healthy controls.

In conclusion, we have identified that a particular subgroup of MCI affects the predictive performance of AD diagnostic models using primary care routine clinical variables. This subgroup (MCI-AD) was characteristically the most similar to AD and, in line with this, were significantly likely to progress to AD over time relative to the other MCI subgroups. These findings suggest that MCI-AD cases are likely representative of patients with prodromal AD. This work highlights the importance of AD diagnostic models focusing specifically on differentiating MCI cases from prodromal AD cases (who are also diagnosed as MCI) as the main way to improve their diagnostic predictive power and translatability.

## Methods

### Data and Patients from the ADNI Cohort

This retrospective study used data generated from the ADNI cohort, publicly available at https://ida.loni.usc.edu/ using data downloaded in April 2024. Patients in the ADNI cohort were diagnosed as either cognitively healthy, MCI, or AD based on the presence of subjective memory complaints, an MMSE score, and CDR ^72^. Healthy controls had no subjective memory complaints, MMSE range of 24-30, and CDR of 0. Patients with MCI had subjective memory complaints, MMSE 24-30 and CDR of 0.5. AD patients similarly had subjective memory complaints, a lower MMSE of 20-26, CDR of greater than 0.5, and meet the criteria for probable AD based on the National Institute of Neurological and Communicative Disorders and Stroke–Alzheimer’s Disease and Related Disorders Association (NINCDS-ADRDA) criteria ^72^. Supplementary Table 1 shows the demographic characteristics of this cohort. We included continuous (e.g. weight, height, BMI, routine blood tests) and categorical (e.g. APOE genotype, physical exam) variables obtained at patients’ baseline visits that we determined would be appropriate for a primary care physician to complete (Supplementary Table 1). We also created additional variables based on cardiovascular metrics including pulse and blood pressure. These included augmented pulse pressure *(systolic BP - diastolic BP) * ls*, pulse to pressure ratio 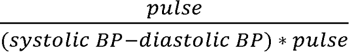, heart stress index 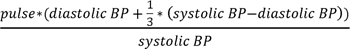, mean arterial pressure 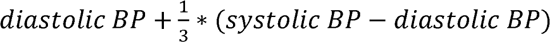, and systolic blood pressure to diastolic blood pressure ratio 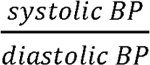. The ADNI cohort study was approved by the institutional review boards of the participating ADNI centers, and all patients provided informed consent.

### Statistical Analyses and Machine Learning

In our initial experiments, all 120 clinical variables (Supplementary Table 1) were used as features in our multi-class diagnostic model. We also performed a filter-based feature selection method to determine the clinical variables that were highly associated with the outcome (i.e. diagnosis). Here, we used a Kruskal-Wallis test (*p* < 0.01) for identifying significant continuous features and Cramer’s V (> 0.11) for categorical ones.

To evaluate the diagnostic potential of the clinical variables, we used CatBoost ^73^, a gradient boosting framework optimized for both categorical and continuous variables. The complete dataset was initially split randomly into an 80% training set and a 20% held-out testing set to evaluate the final models’ performance. To address class imbalances across healthy control, MCI, and AD patients, we performed oversampling using the “imblearn” library ^74^ where the underrepresented class was randomly resampled. During model development, the training set was further randomly subdivided into training and validation datasets using stratified k-fold cross-validation with four folds to ensure balanced representation across classes and mitigate potential biases. Hyperparameter fine-tuning was performed using the “optuna” library ^75^, which employs a Bayesian optimization approach to efficiently search for the optimal combination of parameters. These included the following, with ranges in brackets: maximum number of trees that can be built (iterations; 100-2000), learning rate (0.01-0.3), coefficient at the L2 regularization term of the cost function (1-10), Bayesian bootstrap parameter (0.5-10.0), randomness for scoring splits (1.0-3.0), depth of the trees (3-10), minimum number of training samples in a leaf (1-100), and percentage of features to use at each split selection (0.5-1.0).

To identify the clustering across healthy controls, MCI, and AD patients, we first performed a principal component analysis (PCA) using the filter-based selected features. We then identified subclusters (subgroups) of MCI patients by taking principal component (PC) 1 and PC2 and using Gaussian mixture models for clustering. We confirmed these results using a hierarchical cluster dendrogram. All experiments and optimizations were conducted using python (v.3.11.7) with the libraries listed above and “pandas”, “numpy”, “matplotlib”, “seaborn”, “sklearn”, “catboost”, and “scipy” and Google Colab’s GPU-accelerated environment, which facilitated faster model training and evaluation. Source code is available at https://github.com/Art83/adni_mci.

### Model evaluation

Performance of the machine learning models in this study were evaluated using a 20% held-out testing dataset. For all models, we report performance metrics including sensitivity (correctly identified positive cases), specificity (correctly identified negative controls), PPV (or precision; number of positive cases / total number of predicted positive cases (true and false)), negative predictive value (NPV; number of negative cases / total number of predicted negative cases (true and false)), and AUC (ability to distinguish between positive and negative cases). In the current study, we used specificity, NPV, sensitivity, and PPV as the main indicators of model performance. We note that while AUC is a commonly used performance metric in machine learning studies, it only provides a limited insight into model performance ^76^. This is further supported by previous AD predictive models reporting high AUCs but low sensitivities and are therefore unable to identify incident AD cases ^26^. We used a SHAP (Shapley Additive exPlanations) analysis to evaluate the relative contribution of features to our model performance ^77^. This allowed us to identify those specific features that are most likely to act as translational diagnostic variables for primary care physicians. SHAP was done in python (v.3.11.7) using the “shap” library.

## Data Availability

The data used in this study is from the ADNI cohort and is available at https://ida.loni.usc.edu/.

## Code Availability

Source code is available on GitHub at https://github.com/Art83/adni_mci.

## Acknowledgements

The authors are grateful to the Alzheimer’s Disease Neuroimaging Initiative for providing the data and to all the patients and their families for their involvement in the study. This research did not receive any specific grant from funding agencies in the public, commercial, or not-for-profit sectors. C.A.F. receives salary support from the Neil & Norma Hill Foundation, Annemarie & Arturo Gandioli-Fumagalli Foundation, Perpetual Foundation – John Williams Endowment, and the Hillcrest Foundation.

## Author Contributions

C.A.F. and A.S. jointly contributed to the concept and design, interpretation of data and critical review of the manuscript for important intellectual content. A.S. performed the statistical analyses. C.A.F. acquired the data and wrote the manuscript. All authors reviewed the manuscript.

